# Genetic association studies of fibromuscular dysplasia identify new risk loci and shared genetic basis with more common vascular diseases

**DOI:** 10.1101/2020.09.16.20195701

**Authors:** Adrien Georges, Min-Lee Yang, Takiy-Eddine Berrandou, Mark Bakker, Ozan Dikilitas, Soto Romuald Kiando, Lijiang Ma, Benjamin A. Satterfield, Sebanti Sengupta, Mengyao Yu, Jean-François Deleuze, Delia Dupré, Kristina L. Hunker, Sergiy Kyryachenko, Lu Liu, Laurence Amar, Chad M. Brummett, Dawn M. Coleman, Valentina d’Escamard, Peter de Leeuw, Natalia Fendrikova-Mahlay, Daniella Kadian-Dodov, Jun Z. Li, Aurélien Lorthioir, Marco Pappaccogli, Aleksander Prejbisz, Witold Smigielski, James C. Stanley, Matthew Zawistowski, Xiang Zhou, Sebastian Zoellner, FEIRI investigators, International stroke genetics consortium (ISGC) intracranial aneurysm working group, Megastroke, Marc L. De Buyzere, Stéphanie Debette, Piotr Dobrowolski, Wojciech Drygas, Heather L. Gornik, Jeffrey W. Olin, Jerzy Piwonski, Ernst R. Rietzschel, Ynte Ruigrok, Miikka Vikkula, Ewa Warchol Celinska, Andrzej Januszewicz, Iftikhar J. Kullo, Michel Azizi, Xavier Jeunemaitre, Alexandre Persu, Jason C. Kovacic, Santhi K. Ganesh, Nabila Bouatia-Naji

**Affiliations:** Université de Paris, INSERM, Paris Cardiovascular Research Center, F-75006 Paris, France; Division of Cardiovascular Medicine, Department of Internal Medicine, University of Michigan Medical School, Ann Arbor, MI, USA; Department of Human Genetics, University of Michigan Medical School, Ann Arbor, MI, USA; Department of Neurology and Neurosurgery, University Medical Center Utrecht Brain Center, Utrecht University, Utrecht, The Netherlands; Department of Cardiovascular Medicine, Mayo Clinic, Rochester MN 55902 USA; Icahn Institute for Genomics and Multiscale Biology, Icahn School of Medicine at Mount Sinai, New York, NY, USA; Centre National de Recherche en Génomique Humaine, Institut de Génomique, CEA and Fondation Jean Dausset-CEPH, Evry, France; Hypertension Unit, Assistance Publique-Hôpitaux de Paris, Hôpital Européen Georges Pompidou, F-75015 Paris, France; Department of Anesthesiology, Michigan Medicine, University of Michigan, Ann Arbor, MI, USA; Vascular Surgery Section, Department of Surgery, Michigan Medicine, University of Michigan, Ann Arbor, MI, 48109, USA; Cardiovascular Institute, Icahn School of Medicine at Mount Sinai, New York, NY, USA; Department of Internal Medicine, Division of General Internal Medicine, Section Vascular Medicine, Maastricht University Medical Centre, Maastricht University, Maastricht, the Netherlands; CARIM School for Cardiovascular Diseases, Maastricht University Medical Centre, Maastricht University, Maastricht, the Netherlands; Heart and Vascular Institute, Cleveland Clinic, Cleveland, OH, 44195, USA; Zena and Michael A. Wiener Cardiovascular Institute and Marie-Josée and Henry R. Kravis Center for Cardiovascular Health Icahn School of Medicine at Mount Sinai; Division of Cardiology, Cliniques Universitaires Saint-Luc, Université Catholique de Louvain, 1200 Brussels, Belgium; Division of Internal Medicine and Hypertension Unit, Department of Medical Sciences, University of Turin, Turin, Italy; Department of Hypertension, National Institute of Cardiology, Warsaw, Poland; Department of Demography, University of Lodz, Lodz, Poland; Department of Biostatistics and Center for Statistical Genetics, University of Michigan School of Public Health, Ann Arbor, MI, USA; A full list of authors and their affiliations is provided in the supplementary material.; Univ. Lille, Inserm, CHU Lille, Institut Pasteur de Lille, U1167 - RID-AGE - Labex DISTALZ - Risk factors and molecular determinants of aging-related disease, F- 59000 Lille, France; Department of Cardiovascular Diseases, Ghent University and Ghent University Hospital, Ghent, Belgium; Department of Neurology, Bordeaux University Hospital, Inserm U1219, Bordeaux, France; Department of Epidemiology, Cardiovascular Disease Prevention, and Health Promotion, National Institute of Cardiology, Warsaw, Poland; Human Molecular Genetics, de Duve Institute, Université Catholique de Louvain, 1200 Brussels, Belgium; Université de Paris, Inserm, Centre d’Investigation Clinique 1418, F-75006, Paris, France; Department of Genetics, Assistance-Publiques-Hôpitaux de Paris, Hôpital Européen Georges Pompidou, F-75015 Paris France; Pole of Cardiovascular Research, Institut de Recherche Expérimentale et Clinique, Université Catholique de Louvain, 1200 Brussels, Belgium; Victor Chang Cardiac Research Institute, Darlinghurst, Australia; University of New South Wales, Sydney, Australia

## Abstract

Fibromuscular dysplasia (FMD) is an arteriopathy that presents clinically by hypertension and stroke, mostly in early middle-aged women. We report results from the first genome-wide association meta-analysis of FMD including 1962 FMD cases and 7100 controls. We confirmed *PHACTR1* and identified three new loci (*LRP1, ATP2B1*, and *LIMA1)* associated with FMD. Transcriptome-wide association analysis in arteries identified one additional locus (*SLC24A3)*. FMD associated variants were located in arterial-specific enhancers active in vascular smooth muscle cells and fibroblasts. Target genes are broadly involved in mechanisms related to actin cytoskeleton and intracellular calcium homeostasis, central to vascular contraction. Cross-trait linkage disequilibrium analyses identified positive genetic correlations with blood pressure, migraine and intracranial aneurysm, and an inverse correlation with coronary artery disease, independent from the genetics of blood pressure.

## Introduction

Cardiovascular disease (CVD) is the primary cause of mortality in the world. CVD causes ∼ 18 million deaths each year, of which 85% are due to stroke and myocardial infarction (MI)^1^. Renal artery stenosis is a cause of hypertension, a preventable risk factor for stroke and MI. Renovascular hypertension results from numerous factors, which include atherosclerosis or fibromuscular dysplasia (FMD) in ~10% of cases^2^. While atherosclerosis has been widely studied and its genetic architecture has been well defined, little is known about the pathogenesis or genetics of FMD. To date, only *PHACTR1*, a pleiotropic locus involved in the genetic risk of several cardiovascular and neurovascular diseases, has been reported to be associated with FMD^3^.

FMD occurs predominantly in early middle-aged women (mean age at diagnosis 46-53 years)^4^, thus representing a subset of the population where cardiovascular and neurovascular disease present differently depending on sex^5,6^. FMD is an idiopathic, segmental, non-atherosclerotic disease of the arterial walls, leading to stenosis of small and medium-sized arteries, often associated to dissection, aneurysm, and in some cases arterial tortuosity^4,7-9^. The prevalence of FMD is hard to estimate due to the need of comprehensive vascular imaging to make a definitive diagnosis while concurrently excluding the presence of atherosclerotic plaques. Diagnosis is often made incidentally on imaging (∼3-4% in healthy kidney donors^10^), as part of an investigation to elucidate early onset and/or resistant hypertension, following a stroke event or spontaneous coronary artery dissection, a form of acute myocardial infarction associated with female sex as well^11^.

Investigating the genetic basis of FMD has been a challenging endeavour due to two main reasons: *i*) poor recognition from the general medical community of this underestimated cause of arterial stenosis due to its atypical presentation in women with few cardiovascular risk factors, who are theoretically considered as protected from CVD; *ii*) the effort needed to collect large cohorts of patients where adequate imaging through computed tomographic angiography or magnetic resonance angiography was conducted to precisely define the non-atherosclerotic phenotype and provide sufficient power to conduct genetic studies^4^.

Here we report findings from a meta-analysis of six genome-wide association studies (GWAS) from Europe and the United States of America to investigate the genetic basis of FMD in 1962 patients and 7100 controls. We conducted a single nucleotide polymorphism (SNP)-level GWAS, gene-based GWAS, and transcriptome-wide association study (TWAS) in arteries. We confirmed the previously associated locus *PHACTR1* and identified three novel risk loci associated with FMD at the SNP level and several novel genes associated to FMD at the gene or transcriptome level. Through the integration of annotation datasets generated in fibroblasts, smooth muscle and endothelial cells, combined with public resources available in arteries, we prioritized variants and identified target genes in most loci. We found that risk genes for FMD are specifically and consistently expressed in smooth muscle cells, fibroblasts and arterial tissue and are involved in regulatory mechanisms related to actin cytoskeleton and intracellular calcium homeostasis, a mechanism central to vascular contraction. Using linkage disequilibrium score regression, we found an important genetic overlap between FMD and blood pressure, migraine, intracranial aneurysm, coronary artery disease and MI, and demonstrated that genetics of blood pressure is not driving the genetic association or correlation with FMD.

## RESULTS

### Meta-analysis of six genome-wide association studies revealed new risk loci for FMD

We tested ∼6.5 million common genetic variants (MAF>0.01) in 1962 FMD cases and 7100 controls. All studies involved participants of European ancestry and were adjusted for sex, the first five principal components and individual study genomic control. Three loci contained SNPs associated with FMD at the genome-wide significance level (**Supplementary Figure S1, Table 1, Figure 1a**); the previously identified *PHACTR1* locus on chromosome 6 (lead SNP rs9349379, OR=1.39, 95%CI: 1.28-1.51, *P =* 2.0**×**10^−14^), *LRP1* on chromosome 12 (rs11172113, OR=1.31, 95%CI: 1.20-1.43, *P =* 2.6**×**10^−10^) and rs17249754, located 10kb downstream of *ATP2B1* on chromosome 12 as well (OR=1.41, 95%CI: 1.26-1.58, *P =* 5.9**×**10^−9^). A GWAS restricted to multifocal FMD, which is characterized by multiple stenoses and is the major FMD subtype (91%, **Supplementary Table S1**), identified one additional signal in *LIMA1* on chromosome 12 (rs6580732, OR=1.30, 95%CI: 1.19- .41, *P =* 2.2**×**10^−9,^ **Figure 1a, Supplementary Figure S1, Table 1**). Despite mapping to the same chromosome, *LIMA1, LRP1* and *ATP2B1* loci are fully independent and map on positions 50.5, 57.5 and 90.1 megabases on chromosome 12, respectively. A GWAS in women only cases (87% of patients) identified the same four loci as significantly associated with FMD, with comparable effect sizes and levels of significance (**Supplementary Figure S1, Table 1, Supplementary Tables S2-S4**). Given the small sample size (N=247), a GWAS in men only was not conducted.

**Table 1:** FMD associated variants in SNP association analyses. Meta-analysis was performed using inverse variance-weighted method. Heterogeneity between cohorts was tested using Cochran’s Q statistics and was not significant. Chr: chromosome, EA: effect allele, EAF: effect allele frequency, OR: odds ratio

| Lead Variant |  |  |  |  |  | All FMD |  | Multifocal FMD |  | Women FMD |  |
| --- | --- | --- | --- | --- | --- | --- | --- | --- | --- | --- | --- |
|  |  |  |  |  |  | 1962 cases<br>7100 controls |  | 1578 cases<br>7100 controls |  | 1715 cases<br>5724 controls |  |
| rsID | Chr | Position<br>(hg19) | EA | EAF | Locus | OR<br>(95% CI) | P-value | OR<br>(95% CI) | P-value | OR<br>(95% CI) | P-value |
| rs9349379 | 6 | 12903957 | A | 0.63 | *PHACTR1* | 1.39<br>(1.28 - 1.51) | 2.0×10<sup>-14</sup> | 1.44<br>(1.31 - 1.57) | 5.2×10<sup>-15</sup> | 1.43<br>(1.30 - 1.56) | 1.3×10<sup>-14</sup> |
| rs11172113 | 12 | 57527283 | T | 0.62 | *LRP1* | 1.31<br>(1.20 - 1.43) | 2.6×10<sup>-10</sup> | 1.34<br>(1.22 - 1.46) | 2.0×10<sup>-10</sup> | 1.32<br>(1.20 - 1.44) | 1.5×10<sup>-9</sup> |
| rs17249754 | 12 | 90060586 | G | 0.84 | *ATP2B1* | 1.41<br>(1.26 - 1.58) | 5.9×10<sup>-9</sup> | 1.43<br>(1.26 - 1.62) | 1.7×10<sup>-8</sup> | 1.44<br>(1.27 - 1.63) | 8.1×10<sup>-9</sup> |
| rs6580732 | 12 | 50646279 | T | 0.45 | *LIMA1* | 1.24<br>(1.14 - 1.34) | 1.4×10<sup>-7</sup> | 1.30<br>(1.19 - 1.41) | 2.2×10<sup>-9</sup> | 1.29<br>(1.18 - 1.40) | 7.4×10<sup>-9</sup> |

**Figure 1.**
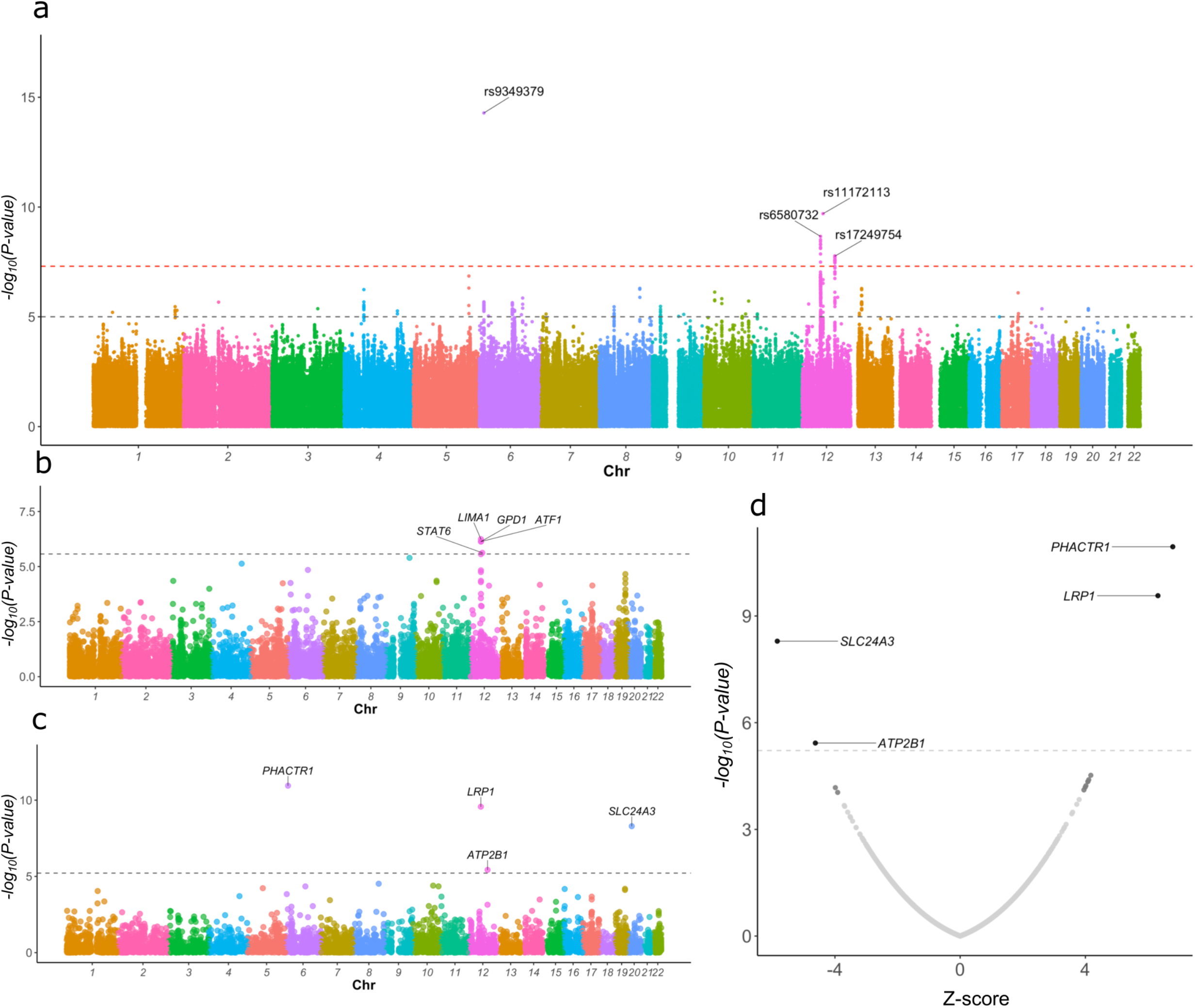
SNP-based, gene-based and transcriptome wide association analyses. **a:** Manhattan plot representation of SNP-based association analysis in multifocal FMD. -log_10_ of association *P*-value is represented on the y-axis, genomic coordinates on the x-axis. Name of lead SNPs with *P*-value ≤ 5**×**10^−8^ are indicated. **b-c:** Manhattan plot representation of **b**: gene-based association analysis in multifocal FMD; **c**: Transcriptome-wide association analysis (TWAS) in FMD with tibial artery gene expression models. -log_10_ of association *P*-value is represented on the y-axis, genomic coordinates on the x-axis. Name of genes with Bonferroni corrected *P*-value ≤ 0.05 are indicated. **d**: Volcano plot representation of FMD TWAS. TWAS Z-score is represented on the x-axis, -log_10_ of TWAS *P*-value on the y-axis. Dashed line represents the threshold for significance adjusted for multiple testing. Name of genes with Bonferroni adjusted *P*-value < 0.05 are indicated.

### FMD associated variants regulate the expression of nearby genes in arterial tissues

To identify potential target genes at FMD associated loci, we queried the GTEx database for eQTL association of lead variants in all tissues available (v8 release, **Figure 2a**)^12^. Interestingly, the lead SNPs in all four loci were identified among top eQTLs of at least one nearby gene in one or more of the three available arterial tissues **(Figure 2a**).

**Figure 2.**
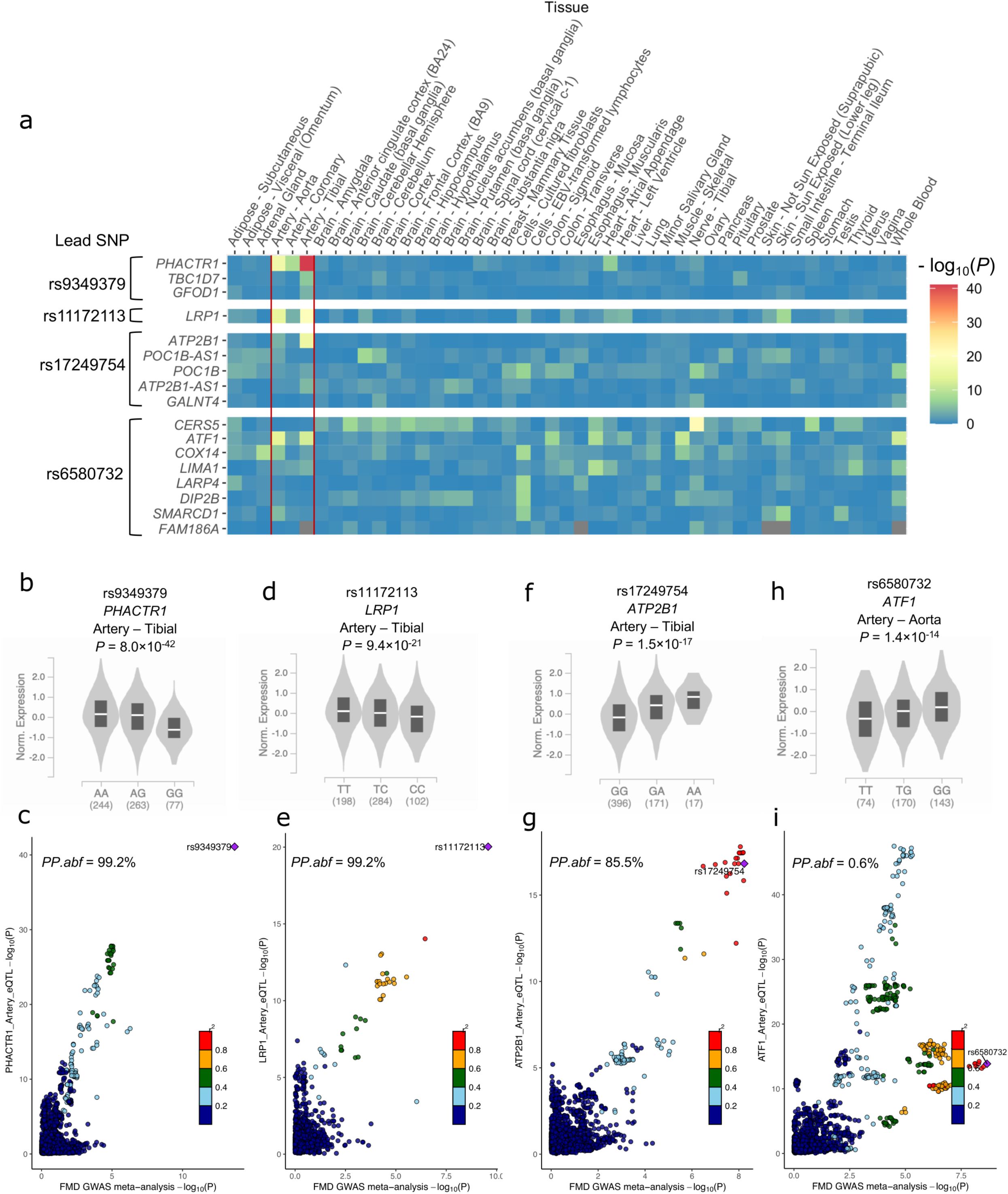
Tissue-wide eQTL signals near FMD loci. **a**: Heatmap representation of eQTL signals at FMD loci. GTEx v8 database was queried for significant eQTLs using the lead variant rsID number. All genes identified as positive eQTL in at least one tissue were selected to calculate eQTL associations in all tissues available in this database. The negative logarithm (-log_10_) of the eQTL association *P*-values are represented in a blue-red colour scale. **b, d, f, h**: Violin plots representing normalized expression of *PHACTR1* (**b**), *LRP1* (**d**), *ATP2B1* (**f**) and *ATF1* (**h**) by genotype of lead SNPs in tibial artery (**b, d, f**) and aortic tissue (**h**). Plots illustrate the best eQTL association in arterial tissue for the lead SNP at each locus. eQTL *P*-value is indicated. **c, e, g, i**: Colocalization plot of FMD association (x-axis, log scale of *P*-value) with arterial eQTL association (y-axis, log-scale of *P*-value) at each locus. Dot colour represents the LD r^2^ with the lead variant in 1000G European samples. FMD lead variant is highlighted (Diamond shape, purple). Approximate Bayes Factor Posterior Probability (*PP*.*abf*) for the two traits to share a common causal variant is indicated.

At the *PHACTR1* locus, FMD risk allele (rs9349379-A) was associated with increased *PHACTR1* expression in all arterial tissues analysed (*P*_*Artery–Tibial*_=8.0**×**10^−42^, **Figure 2b**, *P*_*Artery– Aorta*_=2.0**×**10^−17^, *P*_*Artery–Coronary*_=3.0**×**10^−9^). Colocalization analyses between eQTLs in arterial tissues and FMD association strongly supported rs9349379 to be the causative variant on this locus (posterior probability 99.2%, **Figure 2c**). *PHACTR1* encodes a member of the phosphatase and actin regulator family of proteins implicated in the reorganisation of actin cytoskeleton and tubule formation. Interestingly, rs9349379-*PHACTR1* expression correlation was also found in primary dermal fibroblasts cell lines from FMD patients (N=83) and matched controls (N=70). This finding was observed when all samples were jointly analysed (*P =* 0.01), in the FMD group only (*P =* 0.01), but not in the control group (**Supplementary Figure S2**).

At the *LRP1* locus, rs11172113 was highly correlated with *LRP1* expression in tibial arteries (*P*_*Artery–Tibial*_=9.4**×**10^−21^, **Figure 2d**), aorta (*P*_*Artery–Aorta*_=3.6**×**10^−15^) and to a lesser extent coronary artery tissues (*P*_*Artery–Coronary*_=2.4**×**10^−4^). The FMD risk allele (rs11172113-T) was associated with increased expression of *LRP1* and rs11172113 was also supported by the colocalization analysis as the most likely causal variant at this locus (Posterior probability 99.2%, **Figure 2e**) *LRP1* encodes low density lipoprotein receptor related protein 1, a scavenger receptor involved in numerous cellular processes including intracellular signalling. As for the *ATP2B1* locus, the strongest eQTL association involving rs17249754 in artery tissue was with *ATP2B1* in tibial arteries (*P*_*Artery–Tibial*_=1.5**×**10^−17^, **Figure 2f**) and aorta samples (*P*_*Artery–Aorta*_=5.1**×**10^−5^), whereas no association was detected in coronary arteries. The FMD risk allele (rs17249754-G) was associated with decreased expression of *ATP2B1* in arterial tissues (**Figure 2f)**. Colocalization analysis of FMD association and expression in arterial tissue supports a common causal variant (posterior probability 85.5%, **Figure 2g**). We note that the *ATP2B1* antisense transcript (*ATP2B1-AS)* was also predicted as potentially regulated by the same causal variant (posterior probability 76.6%, **Supplementary Figure S3a**). *ATP2B1* encodes the ATPase plasma membrane Ca^2+^ transporting 1 involved in intracellular calcium homeostasis.

Several eQTL signals were found at the *LIMA1* locus, but none colocalized clearly with the FMD association signal (**Figure 2i, Supplementary Figure S3b-d**). *ATF1* was the strongest eQTL protein-coding gene in arterial tissues (*P*_*Artery–Aorta*_=1.4**×**10^−14^, **Figure 2h**, *P*_*Artery– Tibial*_=8.9**×**10^−13^, *P*_*Artery–Coronary*_=1.7**×**10^−3^), with FMD risk allele (rs6580732-T) being associated with lower expression of *ATF1* (**Figure 2h**). However, colocalization analysis of these two signals was inconclusive (posterior probability = 0.6%, **Figure 2i**). *LIMA1* encodes LIM domain and actin binding 1 involved in actin filament depolymerisation while *ATF1* is the gene encoding activating transcription factor 1, an activating transcription factor involved in cell growth and survival.

### Gene-based association and transcriptome-wide association studies identify additional FMD associated genes

GWAS efficiently captures the effect of single variants affecting traits or diseases but may not detect the combined effect of multiple variants independently affecting disease outcome and located in the same gene. To address this limitation in our genetic investigation, we performed gene-based association analyses using MAGMA^13^, a regression-based gene and gene-set analyses for GWAS data implemented in FUMA^14^. We identified four genes, all on chromosome 12 from two different loci to be associated with FMD in at least one of the tested conditions (all FMD cases, multifocal FMD, women only, Bonferroni corrected *P*<0.05, **Figure 1b, Table 2**). These included three genes at the *LIMA1* locus (*LIMA1, P*_*Women*_=3.0**×**10^−7^, *ATF1, P*_*Multifocal*_=7.3**×**10^−7^ and *GPD1, P*_*Multifocal*_=7.3**×**10^−7^) and *STAT6*, located near *LRP1* (*PMultifocal*=2.4**×**10-6).

**Table 2.**
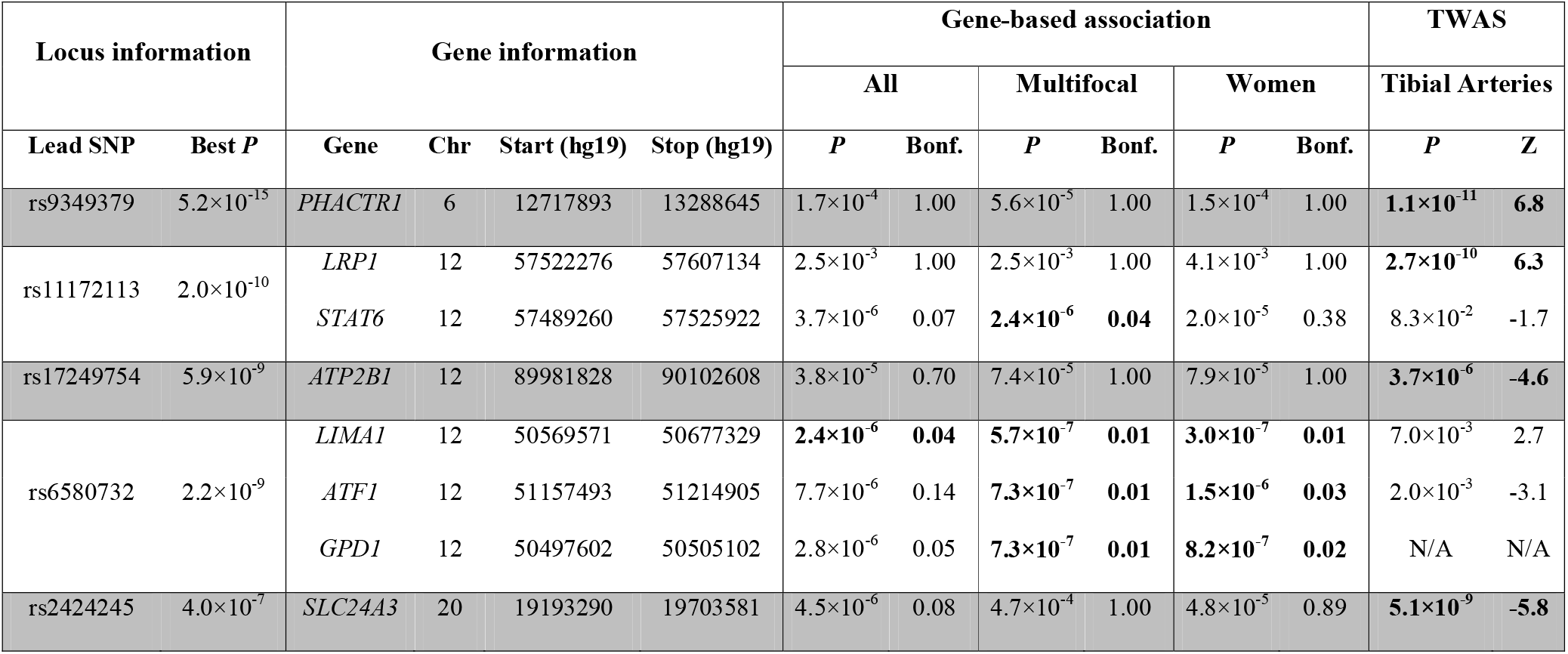
FMD associated genes in gene-based and transcriptome-wide association analyses. The table shows *P*-values (*P*) of gene-based association calculated with MAGMA, uncorrected and corrected for multiple testing, and p-values and Z-scores (Z) of trancriptome-wide association based on gene-expression models from GTEx in tibial artery and GWAS in all FMD. All genes with Bonferroni corrected *P*-value below 0.05 in at least one condition are reported and the condition where the gene association reaches adjusted significance are highlighted in bold. Lead SNP is the one with the lowest *P*-value in GWAS in all FMD. Best *P*: lowest *P*-value of the lead SNP in the three GWAS (all FMD cases, multifocal FMD or women) Chr: Chromosome, Bonf. : Bonferroni corrected *P*-value, N/A: not available

To get a global view of the potential transcriptional effects of FMD associated variants in arteries, we conducted a TWAS using the FUSION software^15^, and gene expression models calculated from tibial artery eQTL analysis from GTEx (v7 release). In line with FMD association and eQTLs analyses, we found a significant association between genetically predicted expression levels of *PHACTR1 (P =* 1.1**×**10^−11^), *LRP1* (*P* = 2.7**×**10^−10^) and *ATP2B1* (*P* = 3.7**×**10^−6^) in arterial tissue and FMD (**Table 2, Figure 1c-d, Supplementary Table S4**). No gene in the *LIMA1* locus was identified as a TWAS hit, although genetically predicted expression of *ATF1* and *LIMA1* were suggestively associated with FMD (*P*_*ATF1*_ = 2**×**10^−3^, *P*_*LIMA1*_ = 7**×**10^−3^, **Table 2**).

In addition to genes located in close proximity of genome-wide FMD-associated loci, we found *SLC24A3* (chromosome 20) to be a robust TWAS hit in tibial artery samples (*P =* 5.1**×**10^−9^, **Figure 1c-d**). Of note, *SLC24A3* was also a suggestive hit in the gene-based association study (*P*_*All*_=4.5**×**10^−6^) (**Table 2**). *SLC24A3* overlaps two independent and suggestive FMD association signals (lead SNP: rs2424245, *P* = 4.0**×**10^−7^, secondary SNP: rs6046121, *P* = 1.2**×**10^−5^, correlation r^2^<0.01, **Supplementary Figure S4**). FMD risk alleles of both variants associated with lower *SLC24A3* expression in artery tissue (rs6046121-A: *P*_*Tibial artery*_ = 2.0**×**10^−11^, *P*_*Aorta*_ = 5.6**×**10^−6^; rs2424245-T: *P*_*Tibial artery*_ = 1.6**×**10^−4^, **Supplementary Figure S4**). Similar to *ATP2B1, SLC24A3* encodes a plasma membrane calcium exchanger also involved in intracellular calcium homeostasis. Results obtained from TWAS using multifocal FMD and women FMD are reported in **Supplementary Figure S5** with overall comparable findings to the whole sample findings.

### FMD associated genes are expressed in vascular smooth muscle cells and fibroblasts

We looked-up the expression of FMD associated genes (**Table 2**) in publicly available mouse aorta single cell RNA-Seq data^16^, 665 tibial artery RNA-Seq samples from GTEx and 153 primary dermal fibroblasts cell lines derived from FMD patients (N = 83) and matched controls (N = 70). Mouse aorta single cell experiment shows that *Phactr1, Lrp1, Atp2b1, Atf1, Lima1, Slc24a3* and *Stat6* were mostly detected in vascular smooth muscle cells (VSMC) and *Phactr1, Lrp1, Atp2b1, Atf1, Lima1* and *Stat6* in fibroblasts clusters, whereas only *Atp2b1* and *Lima1* showed strong presence in endothelial cells (EC) clusters (**Supplementary Figure S6**).

On the other hand, expressions of all the FMD-associated genes were detected in human tibial artery samples from GTEx, although *GPD1* had very low and variable expression levels compared to the other genes (**Supplementary Figure S7**). Interestingly, *SLC24A3* was less expressed in arterial samples of women (*P =* 1.1**×**10^−5^, **Supplementary Figure S8**), consistent with its predicted decreased expression in FMD risk allele carriers. A trend toward higher expression in women was observed for *PHACTR1* and *ATF1* (*P =* 0.013, **Supplementary Figure S8**), also these were consistent with the direction of effect of the FMD increasing risk alleles for *PHACTR1* but not for *ATF1*. However, we did not observe any differences in the expression of top FMD associated genes between fibroblasts samples derived from an all women group of FMD patients and those from sex-matched controls (**Supplementary Figure S9**).

### FMD associated variants are located in regulatory elements active in arterial tissue and VSMCs

The FMD associated variants identified in this study are all located in non-coding regions, either intronic or intergenic. To obtain insights into their potential regulatory function, we generated open chromatin profiles in human carotid artery-derived primary cells (two VSMC and two EC primary cell lines), human coronary artery derived cells (one VSMC and one EC primary cell line), human dermal (two cell lines) and cardiac fibroblasts (one cell line) using ATAC-Seq. We also reanalysed existing data obtained using ATAC-Seq on healthy coronary arteries^17^. Using a common pipeline analysis, we obtained 177,015 to 196,272 peaks from cultured human VSMCs, 120,577 to 137,779 from ECs and fibroblasts, and 54,622 to 70,855 peaks from human coronary arteries (**Figure 3a)**. Global correlation and principal component analyses showed that open chromatin regions of fibroblasts and VSMCs are closely related, whereas ECs and artery samples form separate clusters (**Figure 3b-c**).

**Figure 3.**
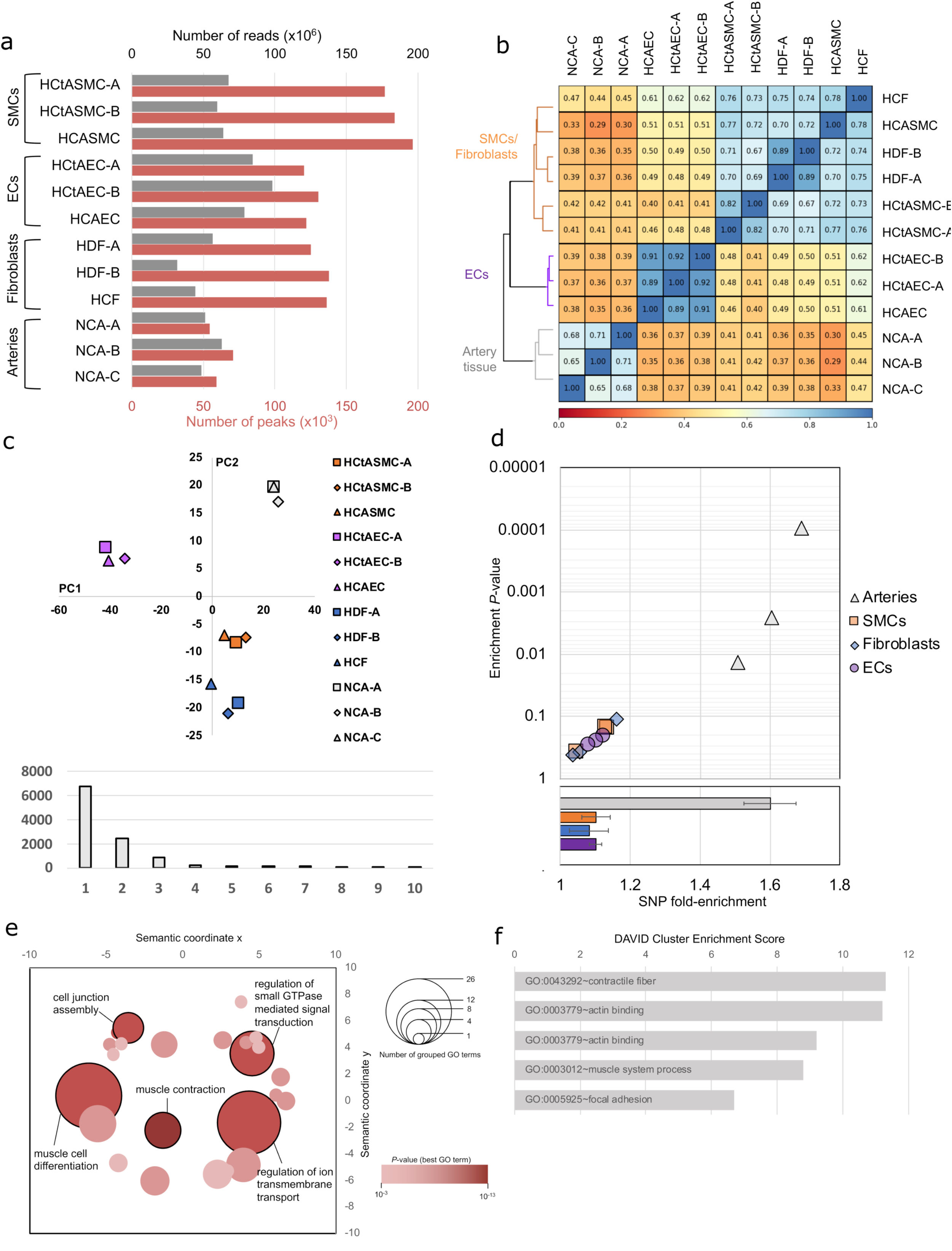
Characterization of open chromatin regions in artery derived primary cells. **a**: Number of reads (grey) and number of peaks (orange) obtained for ATAC-Seq libraries from primary cells and artery tissue. HCtASMC: human carotid artery smooth muscle cells, HCASMC: human coronary artery smooth muscle cells, HCtAEC: human carotid artery endothelial cells, HCAEC: human coronary artery endothelial cells, HDF: human dermal fibroblasts, HCF: human cardiac fibroblasts, NCA: normal coronary arteries. NCA ATAC-Seq libraries were generated and sequenced by Miller and colleagues^17^ and raw reads were retrieved from the sequence read archive (https://www.ncbi.nlm.nih.gov/sra). **b:** Heatmap representation of Spearman correlation and hierarchical clustering of ATAC-Seq datasets. The three main clusters correspond to VSMCs/fibroblasts, ECs and arteries, and are identified on the dendrogram. Rho correlation coefficient is represented by a red-blue colour scale and indicated in each box. **c:** Principal component analysis of ATAC-Seq datasets. Upper panel shows the position of samples with respect to first two principal components. Lower panel indicates the eigenvalues of the first 10 principal components. **d:** Representation of FMD SNPs fold-enrichment (x-axis) and enrichment *P*-value (log scale, y-axis) among indicated ATAC-Seq samples. The overlap of FMD lead SNPs (*P* < 10^−4^ for the lead SNP) and proxies (r^2^ ≥ 0.7) with ATAC-Seq peaks was compared to 500 pools of randomized matched SNPs to calculate the indicated enrichments. Lower panel shows the average fold enrichment in each group of samples. **e:** Bubble graph representing the clustering of enriched (*P* < 10^−3^) gene ontology (GO) Biological Processes terms among genes (N=1425) located in the vicinity of artery-specific open chromatin regions (regions enriched over VSMCs and ECs). Similar terms were grouped using REVIGO webserver (http://revigo.irb.hr/) with “Medium” setting. Bubble size indicates the number of enriched GO terms in each group. Bubble colour indicates the *P*-value of the most enriched term in each group. X-and Y-axes represent arbitrary semantic coordinates. **f**: Bar plot representing the enrichment score of the top 5 clusters obtained by Functional Annotation Clustering of the indicated 1425 genes using DAVID webserver (https://david.ncifcrf.gov/home.jsp). Most enriched term is indicated for each cluster.

Using the GREGOR algorithm^18^, we found that FMD-associated variants were enriched among open chromatin peaks in artery tissue samples (average 1.6-fold enrichment, *P-*values: 9**×**10^−5^-1**×**10^−2^), but not in VSMCs, ECs, or fibroblasts **(Figure 3d)**. We found that, globally, artery-specific ATAC-Seq peaks are overrepresented in the vicinity of genes involved in contractile fibres and muscle system processes (**Figure 3e-f**).

Next, we annotated FMD associated variants with overlapping open chromatin regions we generated in cells and re-analysed in coronary arteries, in addition to histone marks in artery tissue previously generated through the ENCODE project. We defined as potentially causal all variants that overlapped open chromatin peaks in at least one of the above-mentioned arterial tissues or cell types (**Table 3**). Our analyses identified to at least one causal variant per locus. At the *PHACTR1* locus on chromosome 6, the lead variant (rs9349379) belonged to a chromatin region open in arterial tissue but not accessible in VSMCs, fibroblasts or ECs. This variant also overlapped with well-defined enhancer marks in arterial tissue and is strongly supported as the causal variant in this locus (**Figure 4a-b**).

**Table 3.**
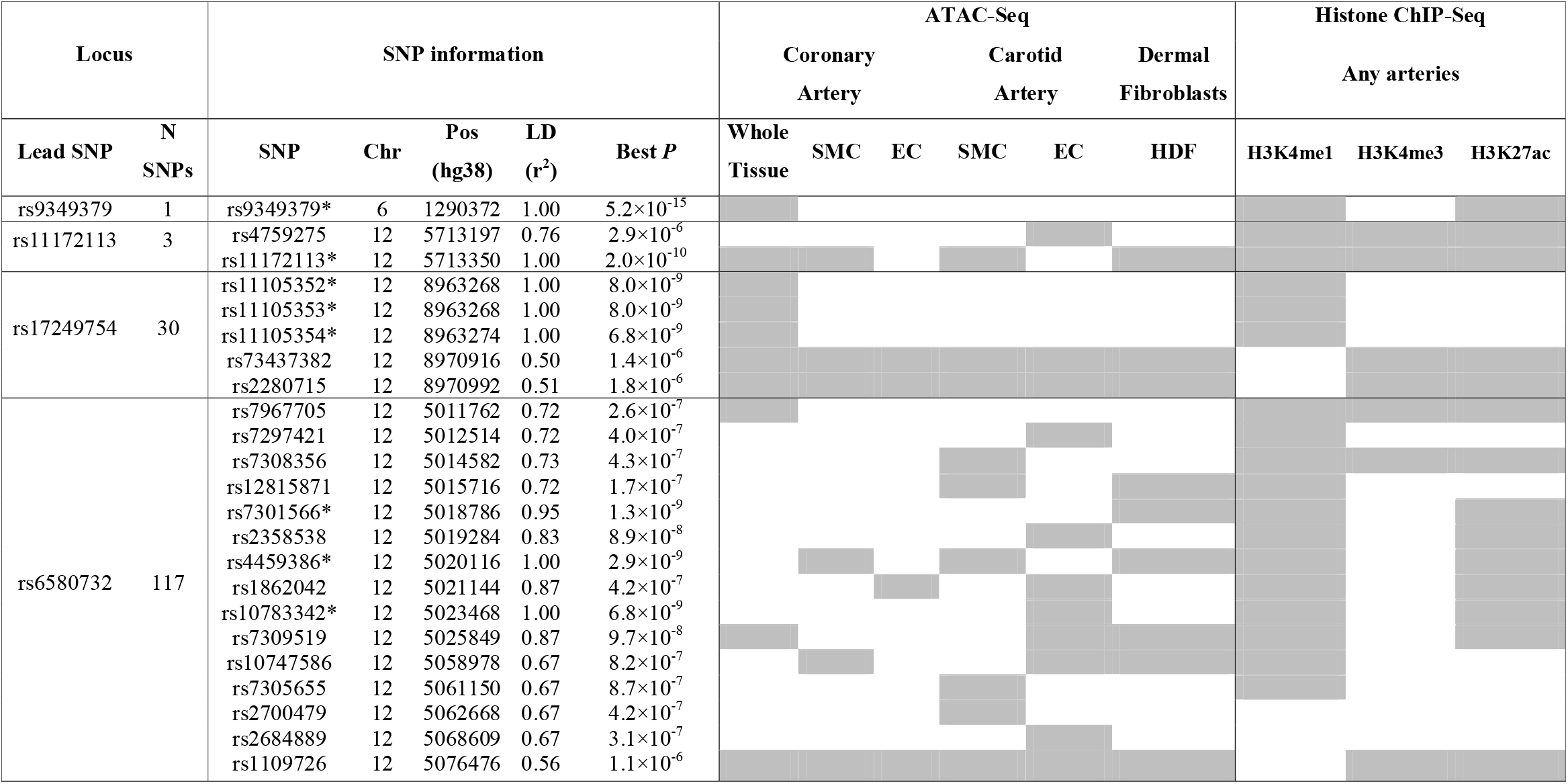
Candidate causal variants at FMD associated loci. All variants with at least suggestive association (P<5×10^−6^) in the global FMD association studies and in high LD (r^2^ ≥ 0.5) with the lead SNP of each locus were tested for overlap with open chromatin regions in coronary artery tissue, carotid/coronary artery derived primary cells, dermal fibroblasts and histone marks in artery tissues (aorta, coronary or tibial arteries) from ENCODE databases. N SNPs indicate the number of SNPs tested at each locus. Best P indicates the lowest *P*-value of the SNP in the three GWAS (all FMD cases, multifocal FMD or women FMD). Grey colour indicates overlap of the variant with at least one dataset of the corresponding category. *: Variant with genome-wide significant association with FMD.

**Figure 4.**
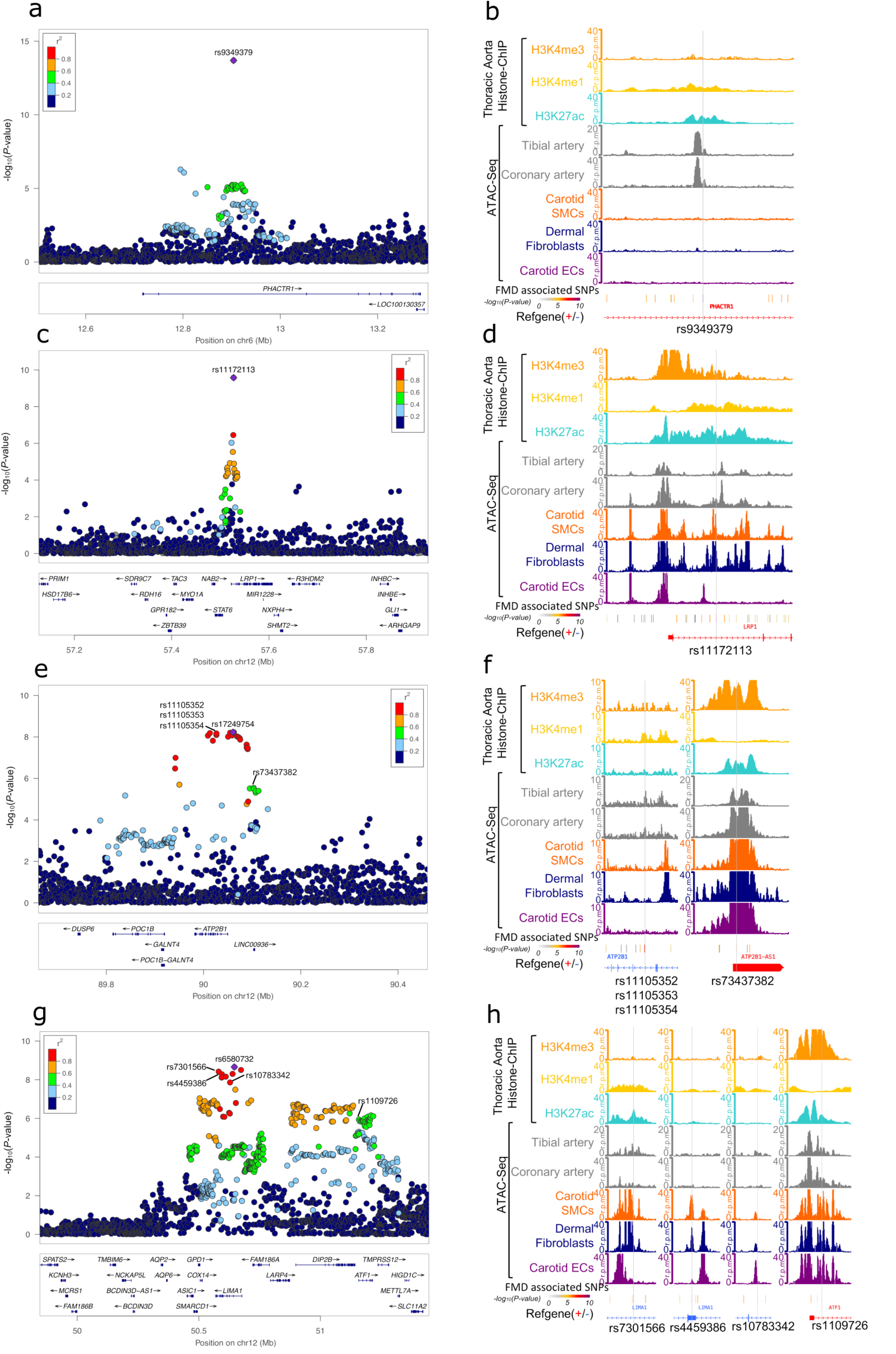
Visualization of potential causal variants genes at FMD-associated loci. **a, c, e, g**: LocusZoom representation of FMD associated loci (**a:** *PHACTR1* locus, **c:** *LRP1* locus, **e**: *ATP2B1* locus, **g**: *LIMA1* locus). Dot colour indicates LD of each variant with the highlighted lead variant (purple diamond). Position and rsID of putative causal variants are indicated. **b, d, f, h**: Genome browser visualization of ATAC-Seq/Histone ChIP read densities (in reads/million, r.p.m.) in the regions surrounding putative causal variants. **b**: *PHACTR1* locus. **d:** *LRP1* locus. **f:** *ATP2B1* locus. **h**: *LIMA1* locus. Grey line highlights variant position.

The lead variant rs11172113 in *LRP1* overlapped with open chromatin peaks in arterial tissue, primary VSMCs and fibroblasts, but not primary ECs (**Figure 4c-d**). Strong enhancer and promoter marks were also present in arterial tissue in this region, which mapped 5kb downstream of *LRP1* promoter.

The *ATP2B1* locus included three highly associated variants (rs11105352, rs11105353 and rs11105354, **Figure 4e**), which all overlapped a small open-chromatin peak specifically observed in arterial tissue, and an enhancer-specific H3K4me1 histone mark (**Figure 4f**). Of note, a suggestively associated SNP (rs73437382, *P* = 2.9×10^−6^), in moderate LD (r^2^ = 0.5) with rs11105354 was located in the promoter sequence of *ATP2B1* and *ATP2B1-AS1* and is thus candidate for causality (**Figure 4e**).

Finally, in the FMD multifocal specific locus *LIMA1*, there were ∼100 tightly correlated variants spanning over 500kb (**Figure 4g**), but only three genome-wide significant variants intronic to *LIMA1* overlapped open chromatin regions (**Figure 4h, Table 3**). rs7301566 overlaps a region active in arteries, VSMCs and fibroblasts and strong enhancer marks in arteries (**Figure 4h**). rs1109726, a suggestively associated variant, shows several marks of regulatory function and maps to the promoter of *ATF1*, ∼480kb downstream of *LIMA1* (**Figure 4h**).

### FMD loci are associated with blood pressure traits but hypertension did not drive genetic association with FMD

FMD diagnosis is often made through radiographic imaging to investigate vascular anomalies in the context of pre-existing and unexplained hypertension, following a stroke event such as due to cervical artery dissection or following MI due to spontaneous coronary artery dissection. To investigate the potentially shared genetic basis of these clinical associations, we curated previously published GWAS on diseases of interest, mainly from GWAS catalog and the UK Biobank summary statistics database (http://www.nealelab.is/uk-biobank). We restricted our curation to FMD lead variants and their proxies (r^2^ ≥ 0.5, Europeans from 1000 Genomes phase 3).

Strikingly, all FMD loci were previously associated with at least one trait related to blood pressure, with the same alleles being associated with increased FMD risk and higher blood pressure/hypertension risk (**Figure 5a**). FMD loci were all associated (*P* ≤ 5×10^−8^) with pulse pressure and at least suggestively (*P* ≤ 1×10^−5^) associated with hypertension, systolic blood pressure and diastolic blood pressure. The lead variants in *SLC24A3* that we identified in the gene-based association and TWAS were associated with blood pressure traits as well (**Figure 5a**).

**Figure 5.**
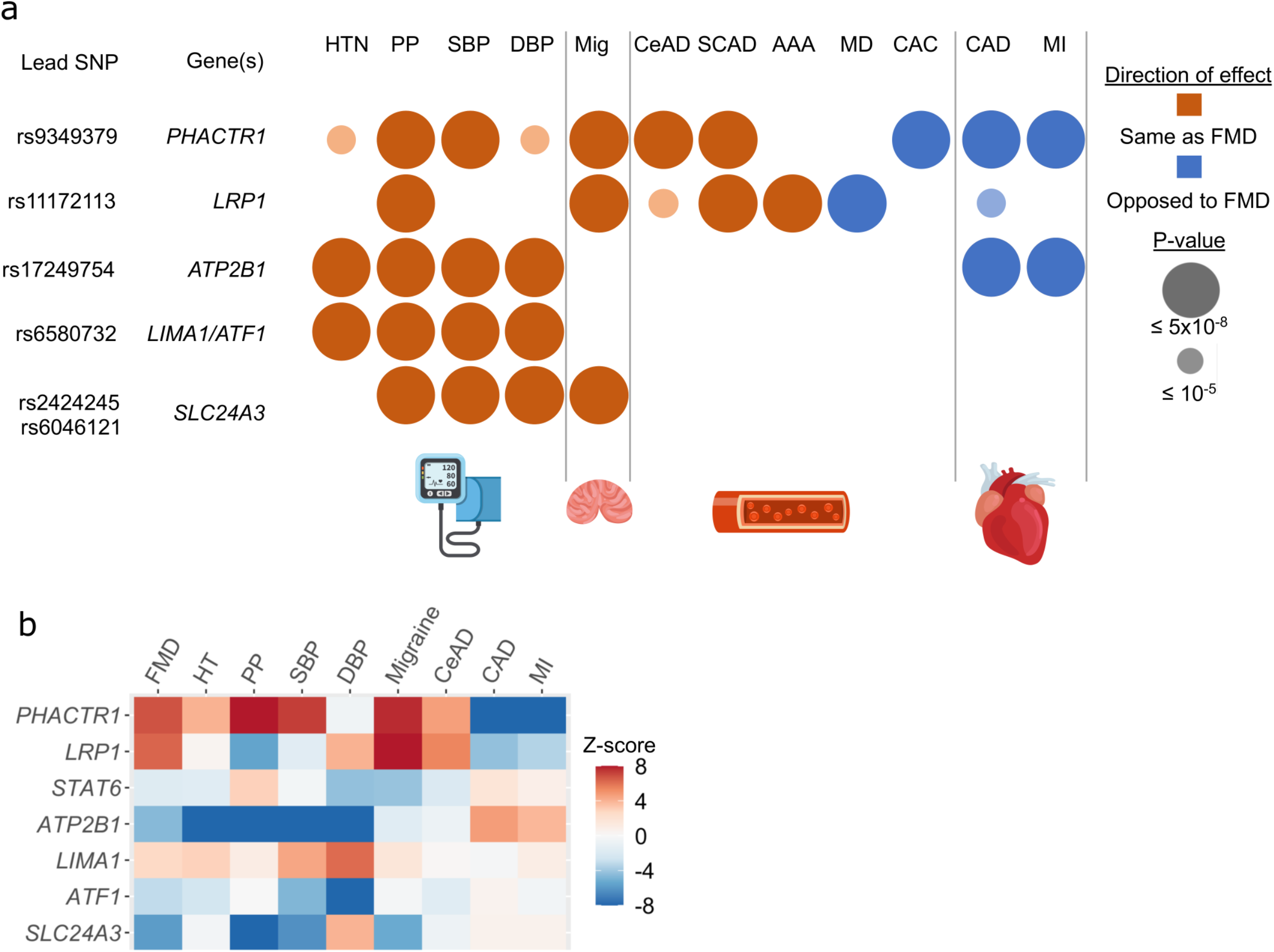
Pairwise trait colocalization of FMD associations. **a**: Associations of FMD loci with other vascular diseases. Variants in LD (r^2^ > 0.5) with the lead SNP were used to query GWAS catalog database (accessed on August 5^th^ 2020), UK BioBank GWAS traits and specific meta-analysis of GWAS for CAD/MI, stroke, blood pressure and intracranial aneurysms. Two independent lead SNPs were retained for *SLC24A3* locus. Overlaps are reported for the following traits/diseases: hypertension (HTN), pulse pressure (PP), systolic blood pressure (SBP), diastolic blood pressure (DBP), migraine (Mig), cervical artery dissection (CeAD), spontaneous coronary artery dissection (SCAD), abdominal aortic aneurysm (AAA), MoyaMoya disease (MD), coronary artery calcification (CAC), coronary artery disease (CAD) and myocardial infarction (MI). Large bubbles indicate association below genome-wide significance for the corresponding trait (*P*<5**×**10^−8^), smaller bubbles correspond to suggestive signals (*P<*1**×**10^−5^). Red colour indicates same direction effects of risk alleles compared to the association with FMD, blue colour opposite direction associations. **b**: Heatmap representation of TWAS Z-Score for FMD associated genes. TWAS was performed with tibial artery gene expression models for the indicated traits or diseases (x-axis). Z-scores are shown for all FMD associated genes in the gene-based and TWAS analyses (y-axis).

Hypertension is reported in a large proportion of patients with FMD in general, and in the majority of FMD cases in this study (51 to 80% of FMD cases, **Supplementary Table S1**). To test for hypertension as a potential confounder driving the observed association signals, we conducted both stratified and adjusted analyses for hypertension status for the GWAS lead variants in the two largest cohorts of our meta-analysis: the French and the UM case control studies (**Supplementary Table S6**). Adjustment for hypertension status marginally modified the effects sizes and level of significance of the associations with FMD at all four loci, including when the association was absent (*LIMA1* in the French study). Given the large proportion of hypertension both in the cases and the controls of the French study, associations with FMD were only observed in the larger stratum of hypertensive patients. However, all four loci showed significant association with FMD both in hypertensive and non-hypertensive individuals in the UM case control study, supporting that these loci impact FMD risk independent of hypertension status in the FMD cases. Concordantly, in the meta-analysis including all 6 individual studies, FMD associations at the top loci remained significant after conditioning FMD association on the genetic association with systolic blood pressure obtained from the recent, large-scale blood pressure GWAS meta-analysis^19^ (**Supplementary Table S7**).

### FMD associated loci have pleiotropic associations with multiple vascular diseases

In addition to blood pressure, we investigated additional associations of the FMD-associated loci and genes with several vascular diseases (**Figure 5a**). *PHACTR1, LRP1* and *SLC24A3* have been previously associated with migraine, with the same alleles at risk for FMD and migraine. *PHACTR1* and *LRP1* were both involved in a wide range of vascular diseases, including cervical artery dissection and spontaneous coronary artery dissection, in addition to abdominal aortic aneurysm for *LRP1. PHACTR1* and *ATP2B1* were associated with coronary artery disease (CAD) and MI, while *LRP1* was suggestively associated to CAD, and in all three loci the risk alleles were the opposite of those associated with FMD. Colocalization of FMD association signals with the most relevant traits, in addition to the comparison of FMD TWAS in tibial arteries with the other diseases (**Figure 5b**), suggested that the same variants cause the associations with FMD, cervical artery dissection, migraine and CAD (**Supplementary Figure S10**).

### Genetic relationships between FMD, cardiovascular and neurovascular diseases

In light of the important number of diseases where FMD loci and genes are involved, we used LD score regression^20^ to calculate the genome-wide correlation between FMD and blood pressure traits, CAD and MI, migraine, several stroke sub-types, lipids and clinical traits related to renal function (**Figure 6a-b, Supplementary Table S8**). FMD was positively correlated with hypertension (r_g_ = 0.37, *P* = 4**×**10^−8^), systolic blood pressure (r_g_ = 0.44, *P* = 5**×**10^−10^), diastolic blood pressure (r_g_ = 0.39, *P* = 2**×**10^−9^) and pulse pressure (r_g_ = 0.36, *P* = 1**×**10^−8^). FMD also correlated positively with migraine (r_g_ = 0.37, *P* = 2**×**10^−4^), intracranial aneurysm (pooled ruptured and unruptured, r_g_ = 0.34, *P* = 7**×**10^−6^), aneurysmal subarachnoid haemorrhage (r_g_ = 0.37, *P* = 4**×**10^−5^), and cervical artery dissection, although this latter did not survive correction for multiple testing (r_g_ = 0.78, *P* = 1**×**10^−2^). FMD was not genetically correlated with several stroke subtypes, CAD and MI. We also found a negative genetic correlation between FMD and low-density lipoprotein cholesterol levels (r_g_ = -0.19, *P* = 1**×**10^−3^). No significant correlation with any of the kidney function related traits was observed (**Figure 6b, Supplementary Table S8**). Interestingly, genetic correlations after conditioning the FMD associations results on genome-wide genetic associations with systolic blood pressure revealed a significant negative genetic correlation with CAD (r_g_ = -0.29, *P* = 6**×**10^−5^) and MI (r_g_ = -0.28, *P =* 2**×**10^−3^) (**Figure 6c, Supplementary Table S9**), indicating that this opposite genetic relation between FMD and CAD/MI is not mediated by common loci between FMD and systolic blood pressure. FMD genetic correlation with migraine was only marginally affected by conditioning on systolic blood pressure genetics (r_g_ = 0.38, *P =* 8**×**10^−5^). On the other hand, the genetic correlations with intracranial aneurysm, subarachnoid haemorrhage and low-density lipoprotein cholesterol levels were less significant after conditioning on systolic blood pressure genetics, indicating that these correlations with FMD are in part due to the common genetic associated loci between FMD and systolic blood pressure (**Figure 6c-d, Supplementary Table S9**). The correlation results were comparable when we conducted the analyses after removing the top FMD associated loci (**Supplementary Tables S10 and S11**).

**Figure 6.**
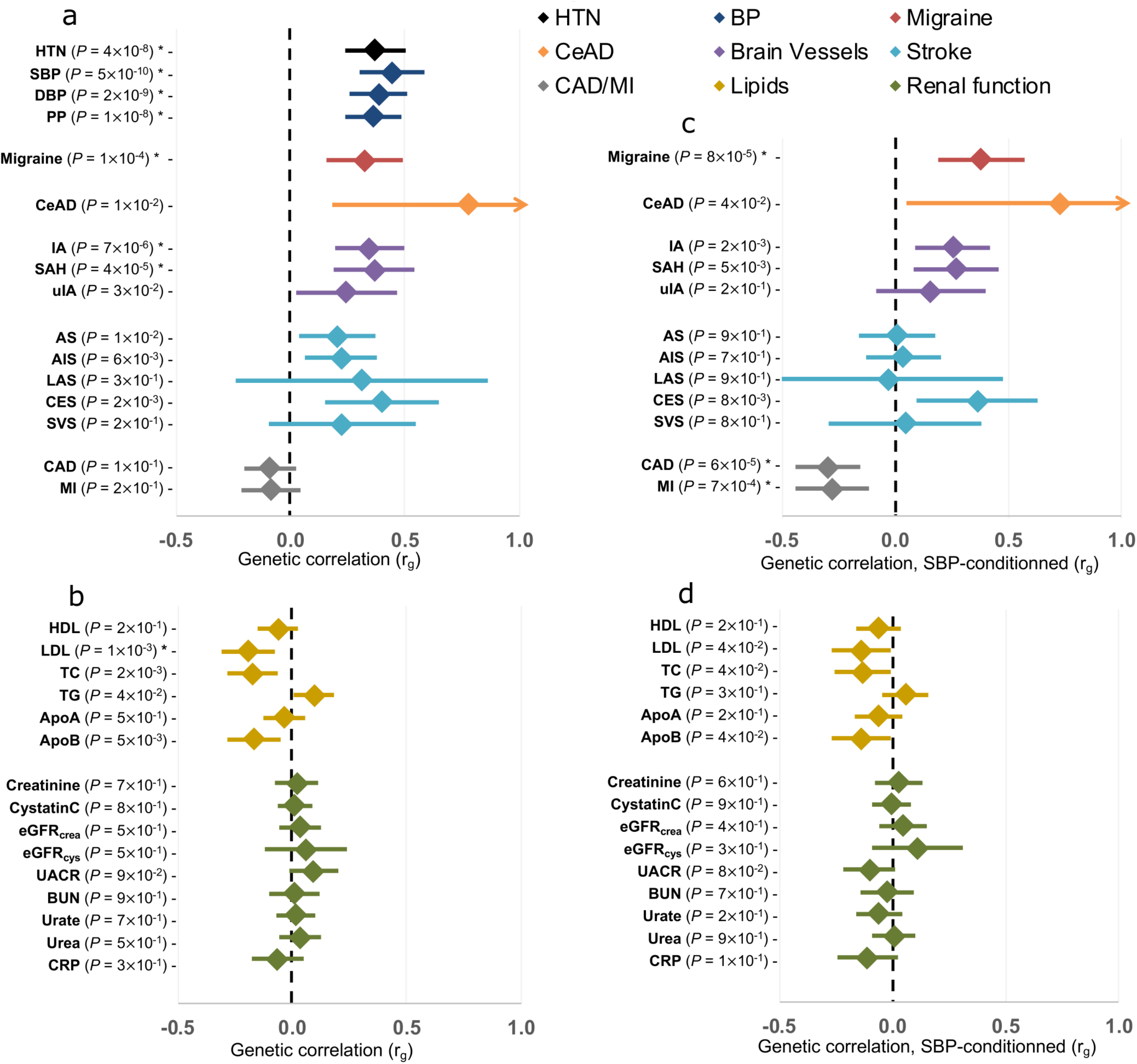
Genetic correlations between FMD and other traits and diseases. **a:** Genetic correlation obtained using LD Score analyses between FMD and vascular diseases and traits. HTN: hypertension, CAD: coronary artery disease, MI: myocardial infarction, AS: any stroke, AIS: any ischemic stroke, LAS: large artery stroke, CES: cardioembolic stroke, SVS: small vessel stroke, IA: intracranial aneurysm, all forms, SAH: aneurysmal subarachnoid haemorrhage, i.e. ruptured intracranial aneurysm, uIA: unruptured intracranial aneurysm. **b:** Genetic correlation obtained using LD Score analyses between FMD and vascular diseases and metabolic traits. HDL: high density lipoprotein, LDL: low density lipoprotein, TC: total cholesterol, TG: triglycerides, ApoA: apolipoprotein A, ApoB: apolipoprotein B, eGFR: estimated glomerular filtration rate (calculated with creatinine or cystatin), UACR: urine albumin to creatinine ratio, BUN: blood urea nitrogen, CRP: C-reactive protein. **c, d**: Genetic correlation results after FMD genetic association statistics was conditioned on systolic blood pressure. *: *P*-value is below the multiple testing threshold for significance (1.6**×**10^−3^ for 31 tests).

## DISCUSSION

Our study is the most comprehensive genetic investigation dedicated to FMD, a non-atherosclerotic arterial disease primarily afflicting middle-aged women with few classical cardiovascular risk factors. Key findings from our study include: *i*) Single SNP GWAS, gene-based GWAS and TWAS analyses in arteries identified three novel risk loci for FMD and involved novel genes while confirming *PHACTR1*, the only known risk locus for FMD. *ii*) Through integration of annotation datasets generated inhouse, combined with public resources, we provided detailed prioritization of FMD risk variants and genes. We found that FMD risk genes are consistently and specifically expressed in VSMCs, fibroblasts and arterial tissue and are involved in regulatory mechanisms related to actin cytoskeleton and intracellular calcium homeostasis, a mechanism central to vascular contraction. *iii*) We found an important genetic overlap between FMD and blood pressure, not only for the associated loci and genes but also globally at the GWAS level, but demonstrated that hypertension is not driving the genetic association with FMD. *iv*) We reported shared genetic bases and potentially biological mechanisms with some but not all cardiovascular and neurovascular diseases, which is only partially driven by shared genetic basis with blood pressure.

The genetic investigation of FMD has been inconclusive for decades due to the lack of a clear genetic model. Efforts to establish large cohorts of FMD patients to conduct well-powered genetic studies are very recent and follow the increased awareness about its relatively high prevalence (∼3%) in asymptomatic individuals^21^. FMD patients are commonly middle-aged women, long considered at low risk for the common cardiovascular diseases.

Our strategy combining single SNP and gene-based GWAS and TWAS in arterial tissues identified three novel loci and several novel genes to contribute to FMD genetic risk. Most of these loci were previously shown to be involved in multiple vascular diseases. *LRP1* is a risk locus for pulse pressure^22^, migraine^23^, aortic abdominal aneurysm^24^, and was recently reported for spontaneous coronary artery dissection^25,26^ involving the same risk allele for FMD that correlates with higher gene expression. However, the opposite allele was reported to increase the risk for MoyaMoya disease^27^ and suggestively for CAD^28^. LRP1 plays important roles in multiple cellular processes relevant to FMD such as the remodelling of the extracellular matrix and VSMCs migration^29^. LRP1 function in VSMCs is mediated partly by the modulation of calcium signalling, leading to deficient vasoconstriction in VSMC-specific *Lrp1*-deficient mice^30^.

Interestingly, two of the genes we identified are directly involved in intracellular calcium homeostasis, a highly relevant molecular mechanism to vascular contractility and vasodilation. *ATP2B1* encodes an ATP-dependent calcium channel specialized in the exportation of calcium ions from the cytoplasm to the extracellular space. *ATP2B1* is a well-established hypertension and blood pressure locus, where the same alleles increase the risk for FMD and hypertension (**Figure 4**). Mice lacking *Atp2b1* specifically in VSMCs exhibit hypertension, higher intracellular calcium levels and increased sensitivity to nicardipine, a calcium channel blocker^31,32^. On the other hand, *SLC24A3*, which we identified through the TWAS analyses in tibial arteries to associate with FMD, encodes a transmembrane sodium/potassium/calcium exchanger also involved in calcium homeostasis^33^. The relevance of impaired vasodilation and/or enhanced vasoconstriction in FMD pathogenesis is supported by our recent study where we reported an enrichment among FMD patients for rare loss-of-function mutations in the gene encoding the receptor for prostacyclin, a major vasodilator hormone^34^. In line with these findings, impaired dilation of arteries in response to sublingual glyceryl trinitrate, a proxy for VSMC dysfunction, was reported in FMD patients, including in arterial segments clinically unaffected by the disease^35^.

The role of some novel genes in the pathogenesis of FMD is still unclear. The signal transducer and activator of transcription 6 gene (*STAT6*) is a ubiquitous transcription factor involved in intracellular processes linked to inflammation, although colocalization and eQTL analyses privileged *LRP1* as causal in this locus. *GPD1* encodes a glycerol-3 phosphate dehydrogenase 1 involved in lipid metabolism, which expression is detected at low levels in arteries. In this locus, where effect estimates of associated variants were higher in multifocal FMD and women, *LIMA1* and *ATF1* are two strong biological candidates. LIMA1 role in actin dynamics is compatible with a potential role in maintaining cell shape, a feature lost in FMD affected VSMCs^36^. A study described a rare frameshift variant in *LIMA1* from a family with inherited low LDL cholesterol and resistance in diet induced hypercholesterolemia in *Lima1* deficient mouse^37^. While its specific role in arteries is not known, ATF1 is a transcriptional effector of the cyclic adenosine monophosphate pathway, which plays a key role in the regulation of vascular tone^38^.

Our study provides robust confirmation of the association with FMD of *PHACTR1*, a pleiotropic locus that is involved in a large number of vascular diseases^39,40^. The functional annotation using open chromatin in vascular cells and arterial tissue, the colocalization analyses, TWAS in arteries and eQTL results, including specifically in FMD patients, all point to rs9349379 as a clear regulator of *PHACTR1*, the most likely causal gene in FMD, and probably in the other vascular diseases as well. The previously suggested regulation of the endothelin-1 gene (*EDN1*)^39^ is not supported by our results, or those from consequent works that used an identical approach of iPSC induced endothelial cells^41^ or measured endothelin-1 plasma levels in FMD patients and matched healthy controls^42^. On the other hand, the phosphatase and actin-binding protein encoded by *PHACTR1* regulates actin stress fibre assembly and cell motility^43^, functions highly relevant to the cellular disorganization that characterizes VSMCs in multifocal FMD affected arteries^36^. Further investigation, especially with *in vivo* models, are needed to more precisely define the function of *PHACTR1* in the context of the genetic risk to a diverse panel of cardiovascular and neurovascular diseases.

Through genetic correlation analyses we were able to globally position the genetic basis of FMD among the genetics of more commonly studied cardiovascular and neurovascular diseases and traits. We showed that FMD shares a significant proportion of its genetic basis with hypertension and blood pressure related traits, not only for the top associated loci and genes but also genome-wide. FMD is often diagnosed in the context of hypertension, although we have demonstrated that hypertension is not driving the genetic association with FMD in the currently studied cohorts. Blood pressure regulation is highly complex and involves multiple organs^44^. The identification of several genes related to intracellular calcium regulation, and the absence of genetic correlation between FMD and multiple renal function traits, suggests that impaired regulation of vascular tone genetically drives the multiple stenoses phenotype observed in FMD that results in increased blood pressure. Future exploration of more common genetic loci between FMD and blood pressure will certainly enlighten additional mechanisms and potentially point to specific therapeutic targets to be privileged in FMD hypertensive patients.

We showed an expected positive genetic correlation between FMD and migraine, which is reported by 25 to 69% of FMD patients^4,8^, and cervical artery dissection, which occurs in the same cerebrovascular beds affected in FMD (i.e. carotid and vertebral arteries). However, we found little support for shared genetics with ischemic stroke subtypes despite FMD being frequently diagnosed in the context of a stroke event. On the other hand, FMD seems to be more genetically related to intracranial aneurysm and aneurysmal subarachnoid haemorrhage. This type of stroke shares several clinical characteristics with FMD, mainly high proportion of patients < 55 years, association with blood pressure and smoking, and a higher propensity of women among patients^45^. Finally, we observed an inverse association between FMD and CAD/MI both for the top loci and genes, and globally when FMD association is conditioned on systolic blood pressure. The elimination of the presence of atherosclerosis as the cause of stenoses and aneurysms is required for FMD diagnosis, which may have influenced this negative correlation with a disease where atherosclerosis is under-represented. FMD is a disease afflicting young to middle-aged women, who are less prone to develop CAD and MI, which predominantly afflict older men. Whether FMD patients are less likely to develop CAD/MI later in life, compared to patients of similar clinical characteristics is not known. Endogenous or exogenous female hormones are considered to be protective factors from CAD/MI^46^, but are suspected as a potential risk factor in FMD pathogenesis, given the high proportion of women among patients (80 to 90%) and the age of diagnosis of FMD (on average ∼50 years)^4^. Oestrogens stimulate the release of vasodilator mediators such as nitric oxide and prostacyclin and inhibit the potent vasoconstrictor endothelin-1^46^. Our study did not point to any direct link with sex hormone metabolism or regulation, except sex differences in the level of expressions among women compared to men for *PHACTR1* and *SLC24A3*, consistent with the direction of effects of FMD risk alleles. More knowledge about detailed biological and physiological roles of both genes are needed to address potential consequences on artery remodelling in sex and atherosclerosis dependent contexts.

In summary, in this first meta-analysis of GWAS for FMD, we report robustly associated loci and genes and provide several new leads toward understanding biological mechanisms of stenosis and dissection in young women in the absence of atherosclerosis. Further investigation of the exact biological effects driven by these genes may shed light on the cause of higher prevalence of FMD in women and provide insights into the shared genetic basis between FMD and more common cardiovascular and neurovascular diseases.

## Data Availability

Data will be fully and freely available after publication acceptance.

## Ethical Statement

All studies involved individual written informed consent from all participants and received approval from respective local ethics committee. The ARCADIA study was approved by the « Comité de Protection des Personnes » CPP Ile-de-France II- ID RCB: 2009-A00288-49. The 3 cities protocol was approved by “comité consultatif de protection des personnes dans la recherche biomédicale Bicêtre Hôpital Bicêtre n°99-28 CCPPRB approved 10/06/99, 11/03/2003 and 17/03/2006. ARCADIA-Pol study was approved by Local Ethics Committee, Institute of Cardiology, IK-NPIA-0021017/1482/17. The WOBASZ II Project was accepted by the Field Bioethics Committee of the Institute of Cardiology in Warsaw (IK-NP-0021-69/1344/12). All centres included in FEIRI received approval from the respective local/ national ethics committees. ASKLEPIOS study was approved by the ethical committee of the Ghent University Hospital, Belgium. The DEFINE case control study is a Mount Sinai Health System Study ID: HSM# 13-00575 / GCO# 13-1118. The Mayo Clinic case control study was approved by Mayo Clinic IRB #08-008355. The UM case control study was approved by University of Michigan IRB #HUM00044507, #HUM00112101 and Cleveland Clinic IRB approval #10-318

## Acknowledgements

We thank all patients who participated in these studies. We thank Dr Antoine Chédid for collecting and managing clinical data of patients in ARCADIA protocol. We thank Patrick Bruneval for his scientific input and exchanges about arterial pathology in FMD. This study contributes to the IdEx Université de Paris ANR-18-IDEX-0001. This work has benefited from the facilities and expertise of the high throughput sequencing core facility of I2BC (Centre de Recherche de Gif – http://www.i2bc.paris-saclay.fr/). ARCADIA-Pol investigators thank Ewa Rudolf, Elzbieta Pazio and Małgorzata Lewandowska, who were responsible for all administrative work. The Steering Committee of the WOBASZ study expresses special thanks for participation in the implementation of the study to: all their co-workers from research teams at six academic centres, to nurses, doctors, and analysts from field research centres located in 16 voivodeships. We acknowledge the Spanish National Cancer Research Centre (CNIO), in the Human Genotyping lab, a member of CeGen where genotyping was performed for part of the cohorts studied. We thank the participants of the study and the Fibromuscular Dysplasia Society of America for facilitating the enrolment of subjects at their annual meetings. We thank the Frankel Cardiovascular Center and M-BRISC program for their support and the University of Michigan Advanced Genomics Core where MGI study performed genotyping. The authors acknowledge the University of Michigan Precision Health Initiative and Medical School Central Biorepository for providing biospecimen storage, management, processing and distribution services and the Center for Statistical Genetics in the Department of Biostatistics at the School of Public Health for genotype data management in support of this research. Some illustrations were designed by macrovector / Freepik.

## Funding

This study was supported by the European Research Council grant (ERC-Stg-ROSALIND-716628) to NB-N and National Institute of Health grant (R01 HL139672) to SKG. The ARCADIA study was sponsored by the Assistance Publique-Hôpitaux de Paris and funded by a grant from the French Ministry of Health (Programme Hospitalier de Recherche Clinique 2009, AOM 08192) and the Fondation de Recherche sur l’Hypertension Artérielle. Genotyping of French study was supported by the French research agency (ANR-13-JSV1-0002) to NB-N. The genotyping of controls from the Three-City Study (3C) was supported by the non-profit organization Fondation Alzheimer (Paris, France) to PA. ARCADIA-Pol study was supported by the grant no. 2.40/III/19 of Institute of Cardiology, Poland. The WOBASZ II Project was financed from the resources at the disposal of the Polish Minister of Health within the framework of the “National Program of Equalization and Accessibility to Cardiovascular Disease Prevention and Treatment for 2010-2012. MV benefited from Fonds de la Recherche Scientifique - FNRS Grant T.0247.19, Belgium. The Spanish National Cancer Research Centre (CNIO), in the Human Genotyping lab, a member of CeGen Biomolecular resources platform (PRB3), is supported by grant PT17 /0019, of the PE I+D+i 2013-2016, funded by *Instituto de Salud Carlos III* and a European regional development fund (ERDF). DEFINE-FMD is supported by NIH grant 1R01HL148167-01A1 to JCK. BAS is supported by the Mayo Clinic Clinician-Investigator Training Program. IJK is additionally supported by NIH grant K24HL137010. The UM study is supported by NHLBI/NIH (R01 HL139672, R01 HL122684), the University of Michigan Taubman Institute, Frankel Cardiovascular Center. S.K.G. is supported by R01HL139672, R01HL122684, and R01HL086694. The Michigan Genomics Initiative was supported by the University of Michigan Precision Health Initiative. The Cleveland Clinic Biorepository was supported by CTSA 1UL1RR024989. The Cleveland Clinic FMD Biorepository has been supported in part by the National Institutes of Health, National Center for Research Resources, CTSA 1UL1RR024989, Cleveland, Ohio. YR received funding from the European Research Council (ERC) under the European Union’s Horizon 2020 research and innovation program (grant agreement No. 852173). Intracranial aneurysm working group acknowledges the support from the Netherlands Cardiovascular Research Initiative: an initiative with support of the Dutch Heart Foundation, CVON2015-08 ERASE.

## Authors contributions

Writing and editing the manuscript: AG, T-EB, SKG, NB-N. Study design / conception: AG, T-EB, SKG, NB-N. Genotyping experiments: J-FD, DD, KLH, MV. Sample /phenotype contribution: LA, CAB, CMB, DMC, HG, SKG, PDL, NF-M, DK-D, JZL, AL, MP, AP, WS, JCS, MZ, XZ, SZ, FEIRI Consortium, PA, MLdB, SD, PD, WD, HLG, JWO, JP, ERR, EW-C, AJ, IJK, MA, XJ, AP, JCK. GWAS analyses: M-LY, T-EB, MB, OD, SRK, LM, BAS, SS, MY, XZ, JZL. Gene-based and LD score analyses: T-EB. Functional annotation experiments: AG, SK, LL. TWAS, in silico functional annotations: AG. eQTL colocalization analyses: M-LY. Data for IA/SAH genetic correlation: MB, ISGC intracranial working group, YR. Data for stroke genetic correlation: MEGASTROKE. eQTL data and analysis in FMD and control fibroblasts: LM, Vd’E, JCK.

## Competing Interest

HLG, SKG, JO, and JCS are non-compensated members of the Medical Advisory Board of the FMD Society of America (FMDSA). SKG is a non-compensated member of the Scientific Advisory Board of SCAD Alliance. Both are non-profit organizations.

## Data availability

Data will be made available after acceptance of the article.

## METHODS

### Patients and control populations

The meta-analysis included participants of European ancestry from six studies: ARCADIA^8^/3C^47^ GWAS, Mayo Vascular Disease Biorepository^48^, DEFINE-FMD study^42^, ARCADIA-POL^49^/WOBASZII^50^ study, University of Michigan/Cleveland Clinic (UM) study^26,51^ and FEIRI^7^/ASKLEPIOS^52^ study. FMD patients presented similar clinical characteristics (**Supplementary Table S1**) and homogeneous diagnosis, exclusion and inclusion criteria. Detailed description of the participating cohorts is available in the supplementary appendix.

### Genome-wide association analyses and meta-analysis

Details on genotyping, variant calling for each cohort and pre-imputation quality control in each study are listed in **Supplementary Table S12**. In brief, genotyping was performed using commercially available arrays. To increase the number of tested SNPs and the overlap of variants available for analysis between different arrays, all European ancestry cohorts imputed genotypes to the most current HRC v1.1 reference panel^53^ on the Michigan Imputation Server^54^. GWAS was conducted in each study under an additive genetic model using PLINK v2.0^55^. Models were adjusted for population structure using the first five principal components, sex (except in the women-only analyses) and study specific genomic control. Prior to meta-analysis, we removed single nucleotide polymorphisms (SNPs) with low minor allele frequencies (MAF) (<□0.01), low imputation quality (r^2^ < 0.8 for French and UM studies and r^2^ <□0.3 for the others studies), and deviations from Hardy-Weinberg equilibrium (*P*□<□10^−^05). A total of 6,477,066 variants met these criteria and were kept in the final results.

Association results were combined using an inverse variance weighted fixed-effects meta-analysis in METAL software^56^, with correction for genomic control. Heterogeneity was assessed using the I^2^ metric from the complete study-level meta-analysis. Between-study heterogeneity was tested using the Cochran Q statistic and considered significant at *P*□≤□10^−3^ Genome-wide significance threshold was set at the level of *P*□=□5.0□×□10^−^08. LocusZoom (http://locuszoom.org/) was used to provide regional visualization of results.

### eQTL and colocalization analyses

We queried GTEx database (v8 release)^57^ with rsID of lead variants at FMD loci in arterial tissues for major associated genes (permutations q-value<0.05). For each identified gene, variant-gene association was queried in all tissues using “eQTL calculator” function on GTEx website (https://gtexportal.org/home/). Uncharacterized non-coding transcripts were excluded from the analysis. For colocalization, three arterial samples (aorta, coronary, tibial) from GTEx were pooled at each locus to compare with the FMD GWAS meta-analysis result and the multifocal FMD GWAS meta-analysis result. We generated colocalization plots using locuscompareR package^58^ and Bayesian posterior probability was calculated using coloc.abf function in R coloc package^59^. The eQTL association results from coronary, tibial and aorta arterial tissues were all retrieved for each transcript at each locus for Bayesian posterior probability analysis, and the minimal *P*-values across these three tissues for each locus were taken for generating the colocalization plots.

### Gene-based and transcriptome-wide association analyses

Gene based association was conducted using the MAGMA tool^13^, implemented in the FUMA platform^14^. Locations of protein-coding genes were defined as the regions from transcription start site to transcription stop site (default option in MAGMA). TWAS was performed using FUSION R/python package^15^. Gene expression models were precomputed from GTEx data (v7 release) and were provided by the authors. Only genes with heritability *P*-value < 0.01 were used in the analysis. Uncharacterized non-coding transcripts were excluded from the analysis. Both tools used linkage disequilibrium information from the European panel of the 1000 Genomes phase 3. Bonferroni multiple testing correction was applied using the p.adjust function in R (v 3.6.1).

### Primary cell culture and ATAC-Seq experiments

With the exception of the dermal fibroblast cell lines obtained under the DEFINE-FMD protocol, other primary cells were purchased from Cell Applications (San Diego, CA) except HDF (ATCC, Manassas, VA) and cultured with 5% CO2 in a 37°C incubator following manufacturer’s instructions. VSMC cells were grown in DMEM supplemented with 5% FBS, Insulin (5µg/mL), EGF (0.5ng/mL), bFGF (2ng/mL) and antibiotics. Cells were at passage 5 (HDF, HCF, HCtAEC, HCAEC) or 6 (HCtASMC, HCASMC) for ATAC-Seq analyses. ATAC-Seq was performed following the Omni-ATAC protocol described previously^60^. Detailed description of ATAC-Seq experiments and analyses is in the supplementary appendix.

### Annotation with epigenomic data

We computed the overlap of variant with open chromatin regions (narrowpeak from MACS2 output + 100bp on each side) and histone-ChIP peaks using bedtools (v2.29.0) annotate function. Full list of peak files used is available in **Supplementary Table S13**. Analysis of SNP enrichment among ATAC-Seq peaks was performed using GREGOR^18^. The lead SNPs from loci associated with *P*-value < 10^−4^ were used as reference for FMD-associated SNPs. We included in the analysis SNPs in LD with lead SNPs (r^2^≥0.7 in the European subset of the 1000 Genomes phase 3 reference panel). We used Integrated Genome Browser (IGB, v9.1.4) to visualize read density profiles and peak positions in the context of human genome^61^.

### Overlap between FMD loci and other traits and diseases

We queried the GWAS catalog database^62^, UK Biobank GWAS summary statistics made publicly available by the Neale lab at the Broad Institute (http://www.nealelab.is/uk-biobank) and GWAS meta-analyses on blood pressure^19^, spontaneous coronary artery dissection^25^, cervical artery dissection^63^, CAD/MI^28^, IA/uIA/SAH (Bakker *et al*., *Nature Genetics*, in press, pre-publication access) and Stroke^64^ with FMD lead SNPs and LD proxies (r^2^ ≥ 0.5 in the European panel of the 1000 Genomes phase 3). We reported vascular phenotypes with at least one variant with genome-wide (*P<*5×10^−8^) or suggestive (*P<*10^−5^) association. Colocalization plots were generated using locuscompareR package^58^. Comparative TWAS in FMD and other traits was performed using HapMap filtered summary statistics (see below).

### Genetic correlation analyses

We used LD score regression to estimate the genetic correlation between FMD and other diseases and traits^65^. Summary statistics were acquired from the respective consortia and are detailed in **Supplementary Table S14**. For each trait, we filtered the summary statistics to the subset of HapMap 3 SNPs to decrease the potential for bias due to poor imputation quality. Correlation analyses were restricted to summary statistics from European ancestry meta-analyses. We used the European LD-score files calculated from the 1000G reference panel and provided by the developers. A *P*<1.6×10^−3^, corresponding to adjustement for 31 independent phenotypes was considered significant. All analyses were performed with the ldsc package (v1.0.1, https://github.com/bulik/ldsc/). We conditioned FMD association on systolic blood pressure genetic association using multi-trait-based conditional and joint analysis (mtCOJO) tool from GCTA pipeline^66^. The resulting summary statistics were then used to calculate genetic correlation between FMD, conditioned on systolic blood pressure, and the previous traits.

## Notes

### Author Declarations

the Comite de Protection des Personnes (CPP) Ile-de-France II- ID RCB: 2009-A00288-49 Comite consultatif de protection des personnes dans la recherche biomedicale Bicetre Hopital Bicetre #99-28 CCPPRB approved 10/06/99, 11/03/2003 and 17/03/2006 Local Ethics Committee, Institute of Cardiology, IK-NPIA-0021017/1482/17 Field Bioethics Committee of the Institute of Cardiology in Warsaw (IK-NP-0021-69/1344/12 Mount Sinai Health System Study ID: HSM# 13-00575 / GCO# 13-1118 Mayo Clinic IRB #08-008355 University of Michigan IRB #HUM00044507, #HUM00112101 and Cleveland Clinic IRB approval #10-318

## REFERENCES

1. World Health, O. World health statistics 2020: monitoring health for the SDGs, sustainable development goals, (World Health Organization, Geneva, 2020).

2. Plouin, P.F. et al. Fibromuscular dysplasia. Orphanet J Rare Dis 2, 28 (2007).

3. Kiando, S.R. et al. PHACTR1 Is a Genetic Susceptibility Locus for Fibromuscular Dysplasia Supporting Its Complex Genetic Pattern of Inheritance. PLoS Genet 12, e1006367 (2016).

4. Gornik, H.L. et al. First International Consensus on the diagnosis and management of fibromuscular dysplasia. Vasc Med 24, 164–189 (2019).

5. Cordonnier, C. et al. Stroke in women - from evidence to inequalities. Nat Rev Neurol 13, 521–532 (2017).

6. Haider, A. et al. Sex and gender in cardiovascular medicine: presentation and outcomes of acute coronary syndrome. Eur Heart J 41, 1328–1336 (2020).

7. Pappaccogli, M. et al. TTHE EUROPEAN/INTERNATIONAL FIBROMUSCULAR DYSPLASIA REGISTRY AND INITIATIVE (FEIRI)- CLINICAL PHENOTYPES AND THEIR PREDICTORS BASED ON A COHORT OF ONE THOUSAND PATIENTS. Cardiovasc Res (2020).

8. Plouin, P.F. et al. High Prevalence of Multiple Arterial Bed Lesions in Patients With Fibromuscular Dysplasia: The ARCADIA Registry (Assessment of Renal and Cervical Artery Dysplasia). Hypertension 70, 652–658 (2017).

9. Olin, J.W. et al. The United States Registry for Fibromuscular Dysplasia: results in the first 447 patients. Circulation 125, 3182–90 (2012).

10. Shivapour, D.M., Erwin, P. & Kim, E. Epidemiology of fibromuscular dysplasia: A review of the literature. Vasc Med 21, 376–81 (2016).

11. Hayes, S.N. et al. Spontaneous Coronary Artery Dissection: Current State of the Science: A Scientific Statement From the American Heart Association. Circulation 137, e523–e557 (2018).

12. The Genotype-Tissue Expression (GTEx) project. Nat Genet 45, 580–5 (2013).

13. de Leeuw, C.A., Mooij, J.M., Heskes, T. & Posthuma, D. MAGMA: generalized gene-set analysis of GWAS data. PLoS Comput Biol 11, e1004219 (2015).

14. Watanabe, K., Taskesen, E., van Bochoven, A. & Posthuma, D. Functional mapping and annotation of genetic associations with FUMA. Nat Commun 8, 1826 (2017).

15. Gusev, A. et al. Integrative approaches for large-scale transcriptome-wide association studies. Nat Genet 48, 245–52 (2016).

16. Kalluri, A.S. et al. Single-Cell Analysis of the Normal Mouse Aorta Reveals Functionally Distinct Endothelial Cell Populations. Circulation 140, 147–163 (2019).

17. Miller, C.L. et al. Integrative functional genomics identifies regulatory mechanisms at coronary artery disease loci. Nat Commun 7, 12092 (2016).

18. Schmidt, E.M. et al. GREGOR: evaluating global enrichment of trait-associated variants in epigenomic features using a systematic, data-driven approach. Bioinformatics 31, 2601–6 (2015).

19. Evangelou, E. et al. Genetic analysis of over 1 million people identifies 535 new loci associated with blood pressure traits. Nat Genet 50, 1412–1425 (2018).

20. Ni, G., Moser, G., Wray, N.R. & Lee, S.H. Estimation of Genetic Correlation via Linkage Disequilibrium Score Regression and Genomic Restricted Maximum Likelihood. Am J Hum Genet 102, 1185–1194 (2018).

21. Hendricks, N.J. et al. Is fibromuscular dysplasia underdiagnosed? A comparison of the prevalence of FMD seen in CORAL trial participants versus a single institution population of renal donor candidates. Vasc Med 19, 363–7 (2014).

22. Giri, A. et al. Trans-ethnic association study of blood pressure determinants in over 750,000 individuals. Nat Genet 51, 51–62 (2019).

23. Gormley, P. et al. Meta-analysis of 375,000 individuals identifies 38 susceptibility loci for migraine. Nat Genet 48, 856–66 (2016).

24. Bown, M.J. et al. Abdominal aortic aneurysm is associated with a variant in low- density lipoprotein receptor-related protein 1. Am J Hum Genet 89, 619–27 (2011).

25. Turley, T.N. et al. Identification of Susceptibility Loci for Spontaneous Coronary Artery Dissection. JAMA Cardiol 5, 1–10 (2020).

26. Saw, J. et al. Chromosome 1q21.2 and additional loci influence risk of spontaneous coronary artery dissection and myocardial infarction. Nat Commun 11, 4432 (2020).

27. Duan, L. et al. Novel Susceptibility Loci for Moyamoya Disease Revealed by a Genome-Wide Association Study. Stroke 49, 11–18 (2018).

28. van der Harst, P. & Verweij, N. Identification of 64 Novel Genetic Loci Provides an Expanded View on the Genetic Architecture of Coronary Artery Disease. Circ Res 122, 433–443 (2018).

29. Bres, E.E. & Faissner, A. Low Density Receptor-Related Protein 1 Interactions With the Extracellular Matrix: More Than Meets the Eye. Front Cell Dev Biol 7, 31 (2019).

30. Au, D.T. et al. LRP1 (Low-Density Lipoprotein Receptor-Related Protein 1) Regulates Smooth Muscle Contractility by Modulating Ca(2+) Signaling and Expression of Cytoskeleton-Related Proteins. Arterioscler Thromb Vasc Biol 38, 2651–2664 (2018).

31. Kobayashi, Y. et al. Mice lacking hypertension candidate gene ATP2B1 in vascular smooth muscle cells show significant blood pressure elevation. Hypertension 59, 854– 60 (2012).

32. Okuyama, Y. et al. The effects of anti-hypertensive drugs and the mechanism of hypertension in vascular smooth muscle cell-specific ATP2B1 knockout mice. Hypertens Res 41, 80–87 (2018).

33. Yang, H. et al. NCKX3 was compensated by calcium transporting genes and bone resorption in a NCKX3 KO mouse model. Mol Cell Endocrinol 454, 93–102 (2017).

34. Georges, A. et al. Rare Loss-of-function Mutations of PTGIR are enriched in Fibromuscular Dysplasia. Cardiovasc Res (2020).

35. Bruno, R.M. et al. Deep Vascular Phenotyping in Patients With Renal Multifocal Fibromuscular Dysplasia. Hypertension 73, 371–378 (2019).

36. Stanley, J.C., Gewertz, B.L., Bove, E.L., Sottiurai, V. & Fry, W.J. Arterial fibrodysplasia. Histopathologic character and current etiologic concepts. Arch Surg 110, 561–6 (1975).

37. Zhang, Y.Y. et al. A LIMA1 variant promotes low plasma LDL cholesterol and decreases intestinal cholesterol absorption. Science 360, 1087–1092 (2018).

38. Morgado, M., Cairrão, E., Santos-Silva, A.J. & Verde, I. Cyclic nucleotide-dependent relaxation pathways in vascular smooth muscle. Cell Mol Life Sci 69, 247–66 (2012).

39. Gupta, R.M. et al. A Genetic Variant Associated with Five Vascular Diseases Is a Distal Regulator of Endothelin-1 Gene Expression. Cell 170, 522-533.e15 (2017).

40. Adlam, D. et al. Association of the PHACTR1/EDN1 Genetic Locus With Spontaneous Coronary Artery Dissection. J Am Coll Cardiol 73, 58–66 (2019).

41. Wang, X. & Musunuru, K. Confirmation of Causal rs9349379- PHACTR1 Expression Quantitative Trait Locus in Human-Induced Pluripotent Stem Cell Endothelial Cells. Circ Genom Precis Med 11, e002327 (2018).

42. Olin, J.W. et al. A Plasma Proteogenomic Signature for Fibromuscular Dysplasia. Cardiovasc Res (2019).

43. Wiezlak, M. et al. G-actin regulates the shuttling and PP1 binding of the RPEL protein Phactr1 to control actomyosin assembly. J Cell Sci 125, 5860–72 (2012).

44. Beevers, G., Lip, G.Y. & O’Brien, E. ABC of hypertension: The pathophysiology of hypertension. Bmj 322, 912–6 (2001).

45. Etminan, N. et al. Worldwide Incidence of Aneurysmal Subarachnoid Hemorrhage According to Region, Time Period, Blood Pressure, and Smoking Prevalence in the Population: A Systematic Review and Meta-analysis. JAMA Neurol 76, 588–597 (2019).

46. Perez-Lopez, F.R., Larrad-Mur, L., Kallen, A., Chedraui, P. & Taylor, H.S. Gender differences in cardiovascular disease: hormonal and biochemical influences. Reprod Sci 17, 511–31 (2010).

47. Vascular factors and risk of dementia: design of the Three-City Study and baseline characteristics of the study population. Neuroepidemiology 22, 316–25 (2003).

48. Ye, Z., Kalloo, F.S., Dalenberg, A.K. & Kullo, I.J. An electronic medical record- linked biorepository to identify novel biomarkers for atherosclerotic cardiovascular disease. Glob Cardiol Sci Pract 2013, 82–90 (2013).

49. Dobrowolski, P. et al. Echocardiographic assessment of left ventricular morphology and function in patients with fibromuscular dysplasia: the ARCADIA-POL study. J Hypertens 36, 1318–1325 (2018).

50. Drygas, W. et al. Multi-centre National Population Health Examination Survey (WOBASZ II study): assumptions, methods, and implementation. Kardiol Pol 74, 681–90 (2016).

51. Fritsche, L.G. et al. Association of Polygenic Risk Scores for Multiple Cancers in a Phenome-wide Study: Results from The Michigan Genomics Initiative. Am J Hum Genet 102, 1048–1061 (2018).

52. Rietzschel, E.R. et al. Rationale, design, methods and baseline characteristics of the Asklepios Study. Eur J Cardiovasc Prev Rehabil 14, 179–91 (2007).

53. Loh, P.R. et al. Reference-based phasing using the Haplotype Reference Consortium panel. Nat Genet 48, 1443–1448 (2016).

54. Das, S. et al. Next-generation genotype imputation service and methods. Nat Genet 48, 1284–1287 (2016).

55. Chang, C.C. et al. Second-generation PLINK: rising to the challenge of larger and richer datasets. Gigascience 4, 7 (2015).

56. Willer, C.J., Li, Y. & Abecasis, G.R. METAL: fast and efficient meta-analysis of genomewide association scans. Bioinformatics 26, 2190–1 (2010).

57. Battle, A., Brown, C.D., Engelhardt, B.E. & Montgomery, S.B. Genetic effects on gene expression across human tissues. Nature 550, 204–213 (2017).

58. Liu, B., Gloudemans, M.J., Rao, A.S., Ingelsson, E. & Montgomery, S.B. Abundant associations with gene expression complicate GWAS follow-up. Nat Genet 51, 768– 769 (2019).

59. Giambartolomei, C. et al. Bayesian test for colocalisation between pairs of genetic association studies using summary statistics. PLoS Genet 10, e1004383 (2014).

60. Corces, M.R. et al. An improved ATAC-seq protocol reduces background and enables interrogation of frozen tissues. Nat Methods 14, 959–962 (2017).

61. Freese, N.H., Norris, D.C. & Loraine, A.E. Integrated genome browser: visual analytics platform for genomics. Bioinformatics 32, 2089–95 (2016).

62. Buniello, A. et al. The NHGRI-EBI GWAS Catalog of published genome-wide association studies, targeted arrays and summary statistics 2019. Nucleic Acids Res 47, D1005–d1012 (2019).

63. Debette, S. et al. Common variation in PHACTR1 is associated with susceptibility to cervical artery dissection. Nat Genet 47, 78–83 (2015).

64. Malik, R. et al. Multiancestry genome-wide association study of 520,000 subjects identifies 32 loci associated with stroke and stroke subtypes. Nat Genet 50, 524–537 (2018).

65. Bulik-Sullivan, B.K. et al. LD Score regression distinguishes confounding from polygenicity in genome-wide association studies. Nat Genet 47, 291–5 (2015).

66. Zhu, Z. et al. Causal associations between risk factors and common diseases inferred from GWAS summary data. Nat Commun 9, 224 (2018).

